# The impact of the COVID-19 pandemic on quality of life, physical and psychosocial wellbeing in breast cancer patients and survivors – a prospective, multicenter cohort study

**DOI:** 10.1101/2020.06.26.20140657

**Authors:** Claudia A Bargon CA, Marilot CT Batenburg, Lilianne E van Stam, Dieuwke R Mink van der Molen, Iris E van Dam, Femke van der Leij, Inge O Baas, Miranda F Ernst, Wiesje Maarse, Nieke Vermulst, Ernst JP Schoenmaeckers, Thijs van Dalen, Rhodé M Bijlsma, Danny A Young-Afat, Annemiek Doeksen, Helena M Verkooijen, UMBRELLA study group

## Abstract

**Purpose:** The COVID-19 pandemic and the resulting social distancing and lockdown measures are having a substantial impact on daily life and medical management of people with breast cancer. We evaluated to what extent these changes have affected quality of life and physical, and psychosocial wellbeing of people (being) treated for breast cancer.

**Methods:** This study was conducted within the prospective Utrecht cohort for Multiple BREast cancer intervention studies and Long-term evaluation (UMBRELLA). Shortly after the implementation of COVID-19 measures, extra questionnaires were sent to 1595 cohort participants, including standard UMBRELLA quality of life (EORTC) questionnaires. Patient-reported outcomes (PROs) were compared to the most recent PROs collected within UMBRELLA before COVID-19. The impact of COVID-19 on PROs was evaluated using mixed models analysis.

**Results:** In total, 1051 patients (66%) completed the questionnaires. One third (n = 327, 31%) reported a higher threshold to contact their general practitioner due to COVID-19. A significant deterioration in emotional functioning was observed (82·6 to 77·9, p < 0.001) and 505 (48%, 95% CI 45-51) patients reported moderate to severe loneliness. Small significant improvements were observed in QoL, physical-, social- and role functioning scores. In the subgroup of 51 patients under active treatment, there was a strong deterioration in social functioning (69·8 to 5·0, p = 0·03).

**Conclusion:** Due to COVID-19, patients (being) treated for breast cancer are less likely to contact physicians, and experience a deterioration in emotional functioning. Patients undergoing active treatment report a strong drop in social functioning. One in two patients reports (severe) loneliness. Online applications facilitating peer contact and e-mental health interventions could support mental health and social interaction times of total lockdown or social distancing.

## INTRODUCTION

With the outbreak of the novel and rapidly spreading coronavirus disease 2019 (COVID-19), many extraordinary emergency measures have been taken in order to prevent and control spread of the virus.^1,2^ National restrictions varied from total lockdown to targeted quarantine and social distancing.^3-5^ Despite drastic efforts, the World Health Organization (WHO) declared the COVID-19 outbreak officially a pandemic on March 11, 2020.^6^

As the COVID-19 pandemic has put health care systems under unprecedented stress, urgent re-arrangements of non COVID-19 related health care has been of vital importance.^5,7,8^ A shift of tasks and responsibilities of health care personnel was needed in order to keep up with the increasing pressure on the health care workforce.^9,10^ To prioritize hospital capacity in both workforce, (intensive care) beds and medical recourses for critically ill COVID-19 patients, elective care was suspended as much as possible while only emergency care and semi-urgent oncological procedures were continued.^5,11^ For breast cancer, surgical procedures were postponed when possible, various types of treatment (chemo- and radiotherapy) were adapted, and follow-up appointments cancelled, postponed or transformed into (video)calls.^8,12^ Also, paramedical (after)care such as medical rehabilitation and psychological support was scaled down to a minimum.^11^

Delays and changes in breast cancer diagnosis, treatment and follow-up protocols due to COVID-19 may induce concerns about recurrence or survival.^12^ This, in combination with concerns about the new viral threat in general, could impair patients’ mental and emotional wellbeing.^13^ Moreover, social support is crucial for supporting quality of life and mental health in people (being) treated for breast cancer.^14,15^ Measures of social distancing or lockdown may interfere with networks of support, and have a negative impact on mental health and emotional functioning.

The purpose of this study was to measure the impact of the COVID-19 pandemic on quality of life, physical and psychosocial functioning of women (being) treated for breast cancer.

## MATERIALS & METHODS

### Study design and participants

The present study was conducted within the ongoing prospective multicenter Utrecht cohort for Multiple BREast cancer intervention studies and Long-term evaLuAtion (UMBRELLA).^16,17^ Since 2013, the UMBRELLA cohort included patients ≥ □ 18 years old, who were referred from six hospitals in the Utrecht region to the Department of Radiation Oncology of the University Medical Center Utrecht (UMCU), the Netherlands. Inclusion criteria were histologically proven invasive breast cancer or ductal carcinoma in situ (DCIS), and the ability to understand the Dutch language (written and spoken). Prior to the first appointment with the radiation oncologist, breast cancer patients were invited to participate in the UMBRELLA cohort. Participants provided informed consent for the longitudinal collection of clinical data and patient-reported outcomes (PROs) through paper or online questionnaires at regular intervals during and after treatment.^16^ Clinical data, including patient, tumor and treatment characteristics, was provided by the Netherlands Cancer Registry (NKR). PROs were collected before the start of radiation therapy (baseline), after three and six months, and each six months up to ten years thereafter through self-reported questionnaires.^16^ The UMBRELLA study adheres to the Dutch law on Medical Research Involving Human Subjects (WMO) and the Declaration of Helsinki (version 2013). The study was approved by the Medical Ethics Committee of the UMCU (NL52651.041.15, METC 15/165) and is registered on clinicaltrials.gov (NCT02839863). This study is reported in accordance with the STrengthening the Reporting of OBservational studies in Epidemiology (STROBE) statement.^18^

### Data collection

Shortly after introduction of the COVID-19 measures in the Netherlands (March 12, 2020), an additional COVID-19 specific online survey was sent out on the 7th of April, 2020, and one reminder was sent on the 15^th^ of April, 2020. The survey included three PRO-questionnaires (the EORTC QLQ-C30/BR23, HADS and the De Jong Gierveld Loneliness Scale) and a COVID-19-specific questions. The survey was conducted among active UMBRELLA cohort participants who were enrolled between October 2013 and April 2020 with a known email address.

The cancer specific Quality of Life core questionnaire (QLQ-C30) of the European Organization for Research and Treatment of Cancer (EORTC) was used to assess global health-related quality of life (QoL), physical functioning, role functioning, cognitive functioning, emotional functioning, social functioning, dyspnea, insomnia, and financial difficulties.^19^ Patients’ future perspective was evaluated with the breast cancer specific module (QLQ-BR23). Each subscale of the EORTC questionnaires includes one to five items, all measured by a 4-point Likert scale.^19^ Global QoL is scored on a 7-point Likert scale.^20^ For each subscale a summary score was calculated according to the EORTC manual.^20^ After linear transformation to a 0 to 100 scale, a higher score represents a better outcome on each domain for functional scales (i.e. QoL, physical-, role-, cognitive-, social-, and emotional functioning), and a lower score represents better outcome for symptom scales (i.e. dyspnea, insomnia, and financial difficulties).^20^

The Hospital Anxiety and Depression Scale (HADS) was used to assess symptoms of anxiety and depression.^21^ The HADS is a 14-item scale and each item has four answer options. Each question scores 0 to 3 points. Patients with scores of 8 or higher are at risk of having anxiety or depressive disorders.^22^

Overall, emotional, and social loneliness was assessed using the 6-item De Jong Gierveld Loneliness Scale.^23^ Each item has three scoring options: “yes”, “more or less”, or “no”. Loneliness scores were calculated using the matching scoring algorithm.^24^ Patients with scores between two and four on the 6-item scale were considered moderately lonely, and patients with a score above four were considered severely lonely were.^24^ Scores above two on each of the 3-item subscales for emotional and social loneliness indicate emotional and/ or social loneliness. ^24^

Additional questions were developed to assess presence of (symptoms resembling) COVID-19 and the impact of COVID-19 on health care consumption and expectations.

PROs during COVID-19 were compared to the most recent pre-COVID-19 questionnaires as obtained within UMBRELLA. We excluded patients from comparative analyses when their most recent questionnaire was completed more than two years before the day that the first COVID-19 patient was diagnosed in the Netherlands (February 27, 2020).

Clinical data was obtained from the electronic patient records and from the UMBRELLA dataset as retrieved from the NKR, and included age at cohort enrolment, Body Mass Index (BMI, calculated with last known mean height and weight), smoking (current, previous, no smoker), self-reported highest educational level (no education, primary school, pre-vocational secondary education, senior general or pre-university secondary education, secondary vocational education, higher professional education, or university degree), surgical treatment, most invasive axillary treatment, (neo)-adjuvant radiation therapy and systemic treatment, currently receiving active treatment, pathological T and N stage (American Joint Committee on Cancer [AJCC] 7^th^ edition).

### Treatment protocols

All UMBRELLA participants underwent mastectomy or breast conserving surgery combined with axillary staging and/or radiotherapy. Before the COVID-19 pandemic, all patients were treated in adherence to the Dutch guidelines for breast cancer treatment, as appropriate.^25^

After the start of the COVID-19 pandemic, modifications to breast cancer treatment protocols were advised by the Dutch Society for Surgical Oncology (NVCO; March 27, 2020), and the Dutch Society for Medical Oncology (NVMO; March 22, 2020). In general, this advice included deferring of surgery in low risk patients, and de-escalation of (neo)adjuvant chemotherapy. Details are presented in Appendix 1. Breast cancer physicians from each of the six referring hospitals (where participants received all breast cancer treatment, except radiotherapy) confirmed to adhere to the new national advice, with the exception of one hospital, which was exclusively reserved for non-COVID-19 care and could therefore continue with standard surgical care without any alterations, while adhering to the new advice regarding medical oncological care and breast reconstructive surgery.

### Statistical analysis

Frequencies, proportions, and means with standard deviations (SD) or medians with interquartile ranges (IQR) as appropriate, were used to describe patient and clinical characteristics, COVID-19 related questions, and PROs.

To measure the impact of COVID-19 on PROs, most recent reported scores from the EORTC-QLQ30/BR23 and the HADS before the start of the COVID-19 pandemic were compared to the PROs during COVID-19. Crude mean EORTC scores were compared with the paired samples t-test and crude median HADS scores with the Wilcoxon signed rank test.

To estimate whether the impact of COVID-19 on clinically relevant PROs varied with time since (active) treatment, participants were categorized into four groups, i.e. active treatment, non-active treatment and enrolled in UMBRELLA < 24 months before the survey, non-active treatment and enrolled 24-60 months before the survey, and non-active treatment and enrolled > 60 months before the survey. A linear mixed effect model for repeated measurements was used to measure the impact of COVID-19 on PROs, and included a patient-specific random intercept, a linear time effect, and an interaction between time since diagnosis and period (pre- /post-COVID-19). To correct for potential confounders, age (linear) was included as fixed variable in the model in the non-actively treated group. In the actively treated group further adjustment was performed for chemotherapy, type of radiotherapy, and type of surgery. Changes in PROs due to COVID-19 were reported as mean differences (MD) with 95% confidence intervals (CI).

All reported p-values were two-sided and p-values < 0·05 were considered statistically significant. Statistical analyses were performed with the use of IBM Statistical Package for Social Sciences (SPSS) software, version 25 (IBM Corp, Armonk, NY).

## RESULTS

Between October 2013 and April 2020, 3239 patients were enrolled in the UMBRELLA cohort (Fig. 1). Of all study participants, 1595 met the inclusion criteria for the present study and were sent the extra COVID-19 survey, of whom 1051 patients (66%) responded. Mean age was 56 years (SD 9.8) and median time since diagnosis was 24 months (IQR 6-42, Table 1). Most patients (56%) were treated for a stage 1 tumor and received breast conserving surgery (77%). Fifty-one participants (4·9%) were receiving active treatment (chemo- and/or radiotherapy) for their breast cancer during the COVID-19 pandemic (Table 2).

**Table 1.**
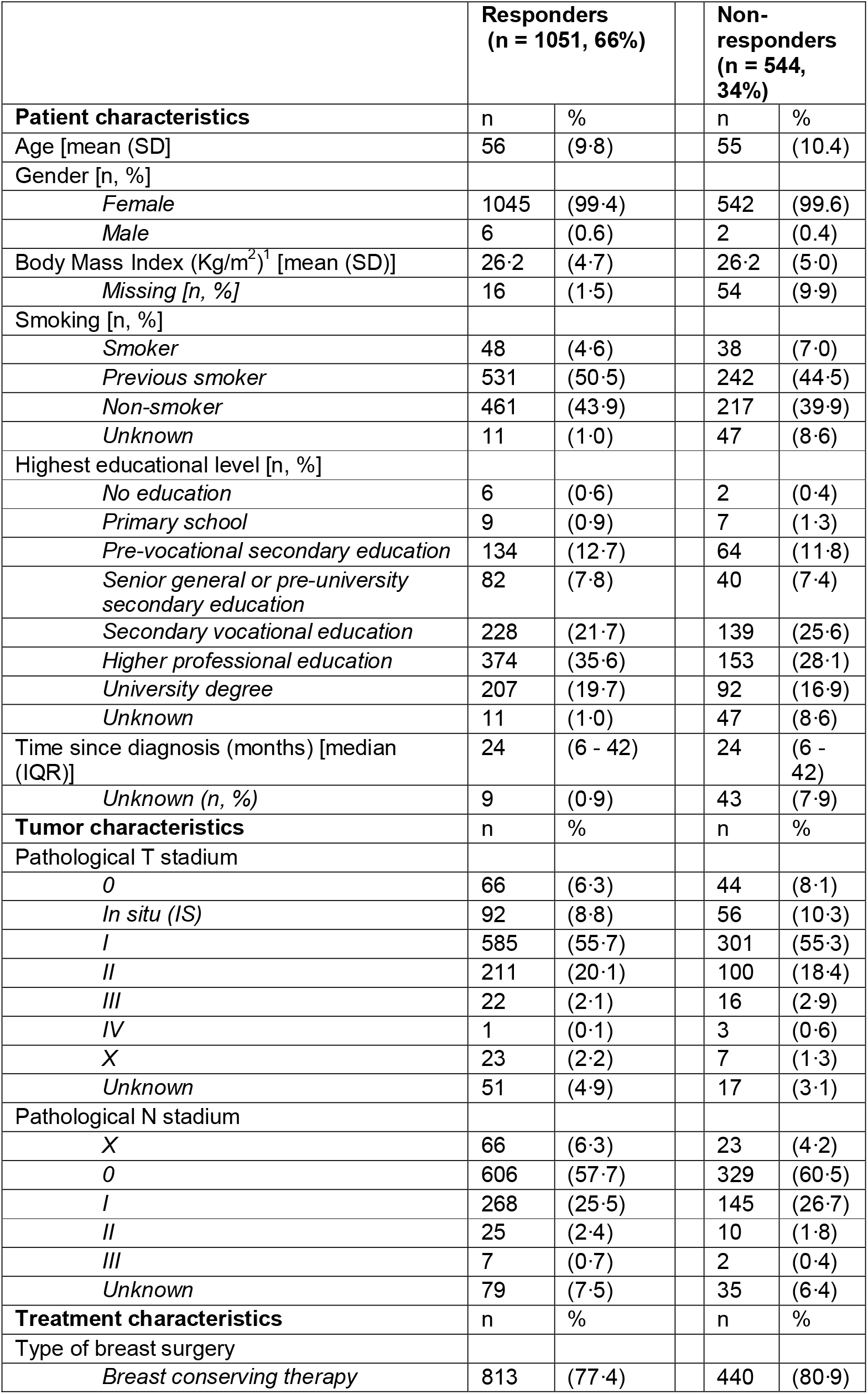
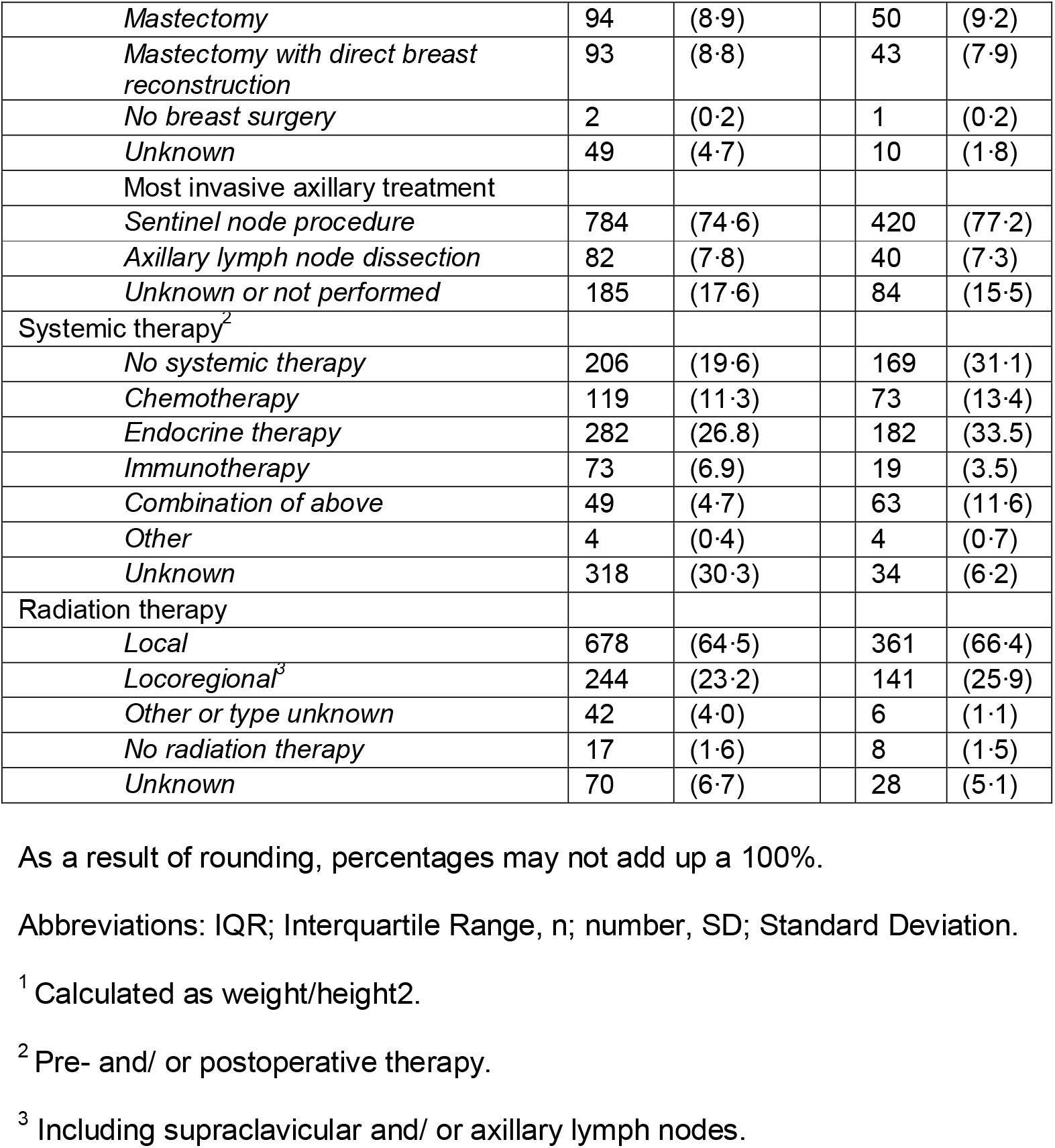
Baseline characteristics of responders and non-responders of the COVID-19 specific online survey that was sent to active UMBRELLA breast cancer cohort participants who were enrolled between October 2013 and April 2020.

|  | Responders<br>(n = 1051, 66%) |  | Non-responders<br>(n = 544, 34%) |  |
| --- | --- | --- | --- | --- |
| **Patient characteristics** | n | % | n | % |
| Age [mean (SD)] | 56 | (9.8) | 55 | (10.4) |
| Gender [n, %] |  |  |  |  |
| *Female* | 1045 | (99.4) | 542 | (99.6) |
| *Male* | 6 | (0.6) | 2 | (0.4) |
| Body Mass Index (Kg/m<sup>2</sup>)<sup>1</sup> [mean (SD)] | 26.2 | (4.7) | 26.2 | (5.0) |
| *Missing [n, %]* | 16 | (1.5) | 54 | (9.9) |
| Smoking [n, %] |  |  |  |  |
| *Smoker* | 48 | (4.6) | 38 | (7.0) |
| *Previous smoker* | 531 | (50.5) | 242 | (44.5) |
| *Non-smoker* | 461 | (43.9) | 217 | (39.9) |
| *Unknown* | 11 | (1.0) | 47 | (8.6) |
| Highest educational level [n, %] |  |  |  |  |
| *No education* | 6 | (0.6) | 2 | (0.4) |
| *Primary school* | 9 | (0.9) | 7 | (1.3) |
| *Pre-vocational secondary education* | 134 | (12.7) | 64 | (11.8) |
| *Senior general or pre-university secondary education* | 82 | (7.8) | 40 | (7.4) |
| *Secondary vocational education* | 228 | (21.7) | 139 | (25.6) |
| *Higher professional education* | 374 | (35.6) | 153 | (28.1) |
| *University degree* | 207 | (19.7) | 92 | (16.9) |
| *Unknown* | 11 | (1.0) | 47 | (8.6) |
| Time since diagnosis (months) [median (IQR)] | 24 | (6 - 42) | 24 | (6 - 42) |
| *Unknown (n, %)* | 9 | (0.9) | 43 | (7.9) |
| **Tumor characteristics** | n | % | n | % |
| Pathological T stadium |  |  |  |  |
| *0* | 66 | (6.3) | 44 | (8.1) |
| *In situ (IS)* | 92 | (8.8) | 56 | (10.3) |
| *I* | 585 | (55.7) | 301 | (55.3) |
| *II* | 211 | (20.1) | 100 | (18.4) |
| *III* | 22 | (2.1) | 16 | (2.9) |
| *IV* | 1 | (0.1) | 3 | (0.6) |
| *X* | 23 | (2.2) | 7 | (1.3) |
| *Unknown* | 51 | (4.9) | 17 | (3.1) |
| Pathological N stadium |  |  |  |  |
| *X* | 66 | (6.3) | 23 | (4.2) |
| *0* | 606 | (57.7) | 329 | (60.5) |
| *I* | 268 | (25.5) | 145 | (26.7) |
| *II* | 25 | (2.4) | 10 | (1.8) |
| *III* | 7 | (0.7) | 2 | (0.4) |
| *Unknown* | 79 | (7.5) | 35 | (6.4) |
| **Treatment characteristics** | n | % | n | % |
| Type of breast surgery |  |  |  |  |
| *Breast conserving therapy* | 813 | (77.4) | 440 | (80.9) |
| *Mastectomy* | 94 | (8.9) | 50 | (9.2) |
| *Mastectomy with direct breast reconstruction* | 93 | (8.8) | 43 | (7.9) |
| *No breast surgery* | 2 | (0.2) | 1 | (0.2) |
| *Unknown* | 49 | (4.7) | 10 | (1.8) |
| Most invasive axillary treatment |  |  |  |  |
| *Sentinel node procedure* | 784 | (74.6) | 420 | (77.2) |
| *Axillary lymph node dissection* | 82 | (7.8) | 40 | (7.3) |
| *Unknown or not performed* | 185 | (17.6) | 84 | (15.5) |
| Systemic therapy<sup>2</sup> |  |  |  |  |
| *No systemic therapy* | 206 | (19.6) | 169 | (31.1) |
| *Chemotherapy* | 119 | (11.3) | 73 | (13.4) |
| *Endocrine therapy* | 282 | (26.8) | 182 | (33.5) |
| *Immunotherapy* | 73 | (6.9) | 19 | (3.5) |
| *Combination of above* | 49 | (4.7) | 63 | (11.6) |
| *Other* | 4 | (0.4) | 4 | (0.7) |
| *Unknown* | 318 | (30.3) | 34 | (6.2) |
| Radiation therapy |  |  |  |  |
| *Local* | 678 | (64.5) | 361 | (66.4) |
| *Locoregional<sup>3</sup>* | 244 | (23.2) | 141 | (25.9) |
| *Other or type unknown* | 42 | (4.0) | 6 | (1.1) |
| *No radiation therapy* | 17 | (1.6) | 8 | (1.5) |
| *Unknown* | 70 | (6.7) | 28 | (5.1) |

As a result of rounding, percentages may not add up a 100%.
Abbreviations: IQR; Interquartile Range, n; number, SD; Standard Deviation.
<sup>1</sup> Calculated as weight/height<sup>2</sup>.
<sup>2</sup> Pre- and/ or postoperative therapy.
<sup>3</sup> Including supraclavicular and/ or axillary lymph nodes.

**Table 2.** COVID-19 specific questions (n = 1051).

|  | Number<br>of<br>patients | Percentage |
| --- | --- | --- |
|  | n | (%) |
| **Are you currently receiving active breast cancer treatment?<sup>1</sup>** |  |  |
| Yes | 51 | (4.9) |
| No | 1000 | (95.1) |
| **Are / were you infected by the COVID-19?** |  |  |
| Yes, confirmed by nasopharyngeal swab | 1 | (0.1) |
| Possibly, I have or had fever | 100 | (9.5) |
| No, I was tested negative | 9 | (0.9) |
| No, I had/ have no symptoms and I was not tested | 941 | (89.5) |
| **Do the current COVID-19 measure affect your current treatment or (after)care?** |  |  |
| Yes | 286 | (27.2) |
| No | 765 | (72.8) |
| **Do you expect that the current COVID-19 measures will affect your treatment or (after)care in the future?** |  |  |
| Yes | 250 | (23.8) |
| No | 801 | (76.2) |
| **Did the threshold to contact your <u>general practitioner</u> change, because of the COVID-19 situation?** |  |  |
| Yes, I contact my general practitioner more easily | 19 | (1.8) |
| Yes, I contact my general practitioner less easily | 327 | (31.1) |
| No | 705 | (67.1) |
| **Did the threshold to contact the <u>physicians treating your breast cancer</u> change, because of the COVID-19 situation?** |  |  |
| Yes, I contact my breast cancer physician(s) more easily | 8 | (0.8) |
| Yes, I contact my breast cancer physician(s) less easily | 162 | (15.4) |
| No | 881 | (83.8) |
| **Did the threshold to discuss your breast cancer diagnosis or breast cancer (treatment) related symptoms with <u>family and friends</u> change, because of the COVID-19 situation?** |  |  |
| Yes, I contact my friends and family more easily | 14 | (1.3) |
| Yes, I contact my friends and family less easily | 87 | (8.3) |
| No | 950 | (90.4) |
| **Are you worried about your financial situation as a result of COVID-19?** |  |  |
| Not at all | 663 | (63.1) |
| A little bit | 320 | (30.4) |
| Quite a bit | 53 | (5.0) |
| Very much | 15 | (1.4) |

As a result of rounding, percentages may not add up a 100%.
<sup>1</sup> Active treatment includes chemotherapy and/ or radiation therapy.

**Figure 1.**
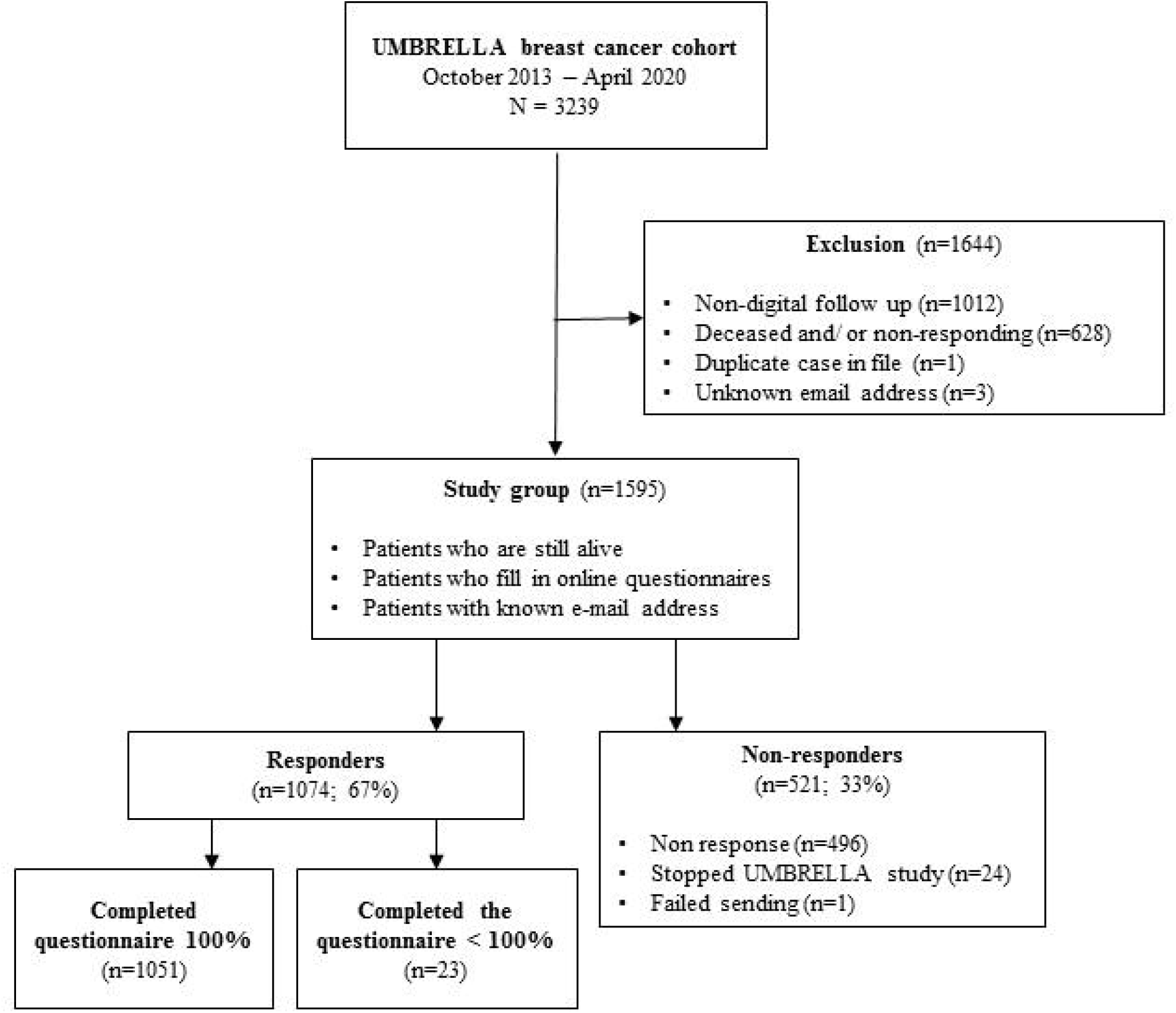
Flowchart of inclusion of study participants from UMBRELLA-cohort. Participants who did not fill out the COVID-19 questionnaire completely (n= 23) were considered non-responders.

### Physical and psychosocial wellbeing during COVID-19

Of all responders, one patient (0·1%) had confirmed COVID-19 infection and 100 patients (9·5%) indicated to have been possibly infected as they experienced symptoms of fever, but they had not been tested for the virus. Twenty-seven percent (n = 286) of all responders felt that the COVID-19 measures affected their current treatment or (after)care, and 24% (n = 250) felt that these measures were likely to affect their (after)care in the future (Table 2).

Almost one third (n = 327, 31%) reported a higher threshold to contact their general practitioner due to the COVID-19 outbreak, and 162 patients (15%) indicated to be less likely to contact the physician treating their breast cancer. Family and friends were contacted less easily by 87 responders (8%). Most responders (n = 983, 95%) were not or a little bit worried about their financial situation as a result of COVID-19 (Table 2).

During the COVID-19 pandemic, 409 of all responders (39%) reported moderate feelings of loneliness, and 96 (9·3%) felt severely lonely (Table 3). Of these, 202 patients (40%) felt socially lonely, and 396 patients (78·4%) felt emotionally lonely.

**Table 3.**
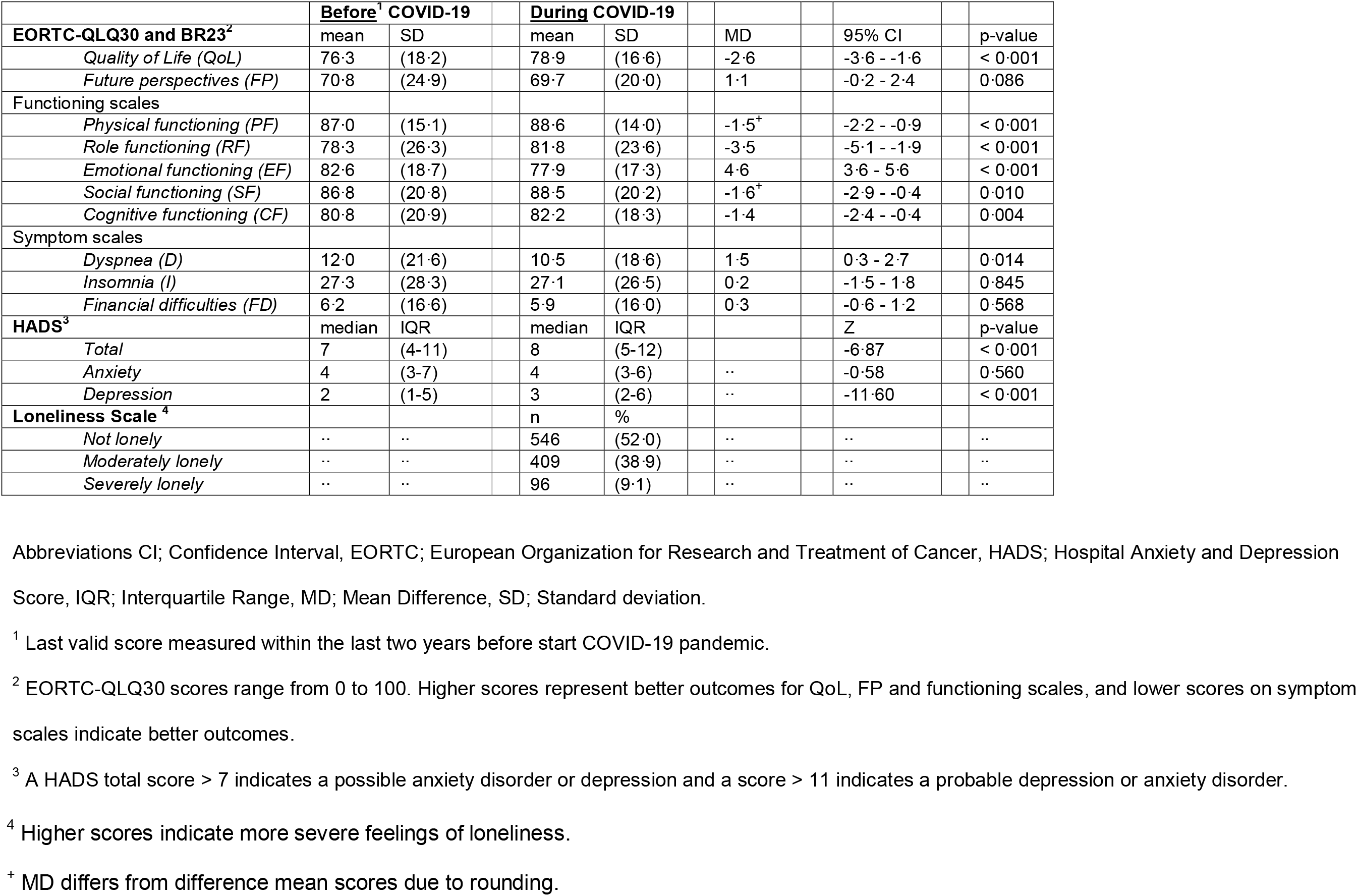
Comparison of crude patient-reported outcomes of all patients who completed the same questionnaire during and 2 years before COVID-19 (n =1022 for EORTC, and n = 942 for HADS). Loneliness score was only measured during COVID-19 (n = 1051).

|  | **Before<sup>1</sup> COVID-19** |  | **During COVID-19** |  |  |  |  |  |
| --- | --- | --- | --- | --- | --- | --- | --- | --- |
| **EORTC-QLQ30 and BR23<sup>2</sup>** | mean | SD | mean | SD | MD |  | 95% CI | p-value |
| *Quality of Life (QoL)* | 76.3 | (18.2) | 78.9 | (16.6) | -2.6 |  | -3.6 - -1.6 | < 0.001 |
| *Future perspectives (FP)* | 70.8 | (24.9) | 69.7 | (20.0) | 1.1 |  | -0.2 - 2.4 | 0.086 |
| Functioning scales |  |  |  |  |  |  |  |  |
| *Physical functioning (PF)* | 87.0 | (15.1) | 88.6 | (14.0) | -1.5<sup>+</sup> |  | -2.2 - -0.9 | < 0.001 |
| *Role functioning (RF)* | 78.3 | (26.3) | 81.8 | (23.6) | -3.5 |  | -5.1 - -1.9 | < 0.001 |
| *Emotional functioning (EF)* | 82.6 | (18.7) | 77.9 | (17.3) | 4.6 |  | 3.6 - 5.6 | < 0.001 |
| *Social functioning (SF)* | 86.8 | (20.8) | 88.5 | (20.2) | -1.6<sup>+</sup> |  | -2.9 - -0.4 | 0.010 |
| *Cognitive functioning (CF)* | 80.8 | (20.9) | 82.2 | (18.3) | -1.4 |  | -2.4 - -0.4 | 0.004 |
| Symptom scales |  |  |  |  |  |  |  |  |
| *Dyspnea (D)* | 12.0 | (21.6) | 10.5 | (18.6) | 1.5 |  | 0.3 - 2.7 | 0.014 |
| *Insomnia (I)* | 27.3 | (28.3) | 27.1 | (26.5) | 0.2 |  | -1.5 - 1.8 | 0.845 |
| *Financial difficulties (FD)* | 6.2 | (16.6) | 5.9 | (16.0) | 0.3 |  | -0.6 - 1.2 | 0.568 |
| **HADS<sup>3</sup>** | median | IQR | median | IQR |  |  | Z | p-value |
| *Total* | 7 | (4-11) | 8 | (5-12) |  |  | -6.87 | < 0.001 |
| *Anxiety* | 4 | (3-7) | 4 | (3-6) | .. |  | -0.58 | 0.560 |
| *Depression* | 2 | (1-5) | 3 | (2-6) | .. |  | -11.60 | < 0.001 |
| **Loneliness Scale<sup>4</sup>** |  |  | n | % |  |  |  |  |
| *Not lonely* | .. | .. | 546 | (52.0) | .. |  | .. | .. |
| *Moderately lonely* | .. | .. | 409 | (38.9) | .. |  | .. | .. |
| *Severely lonely* | .. | .. | 96 | (9.1) | .. |  | .. | .. |

Abbreviations CI; Confidence Interval, EORTC; European Organization for Research and Treatment of Cancer, HADS; Hospital Anxiety and Depression
Score, IQR; Interquartile Range, MD; Mean Difference, SD; Standard deviation.
<sup>1</sup> Last valid score measured within the last two years before start COVID-19 pandemic.
<sup>2</sup> EORTC-QLQ30 scores range from 0 to 100. Higher scores represent better outcomes for QoL, FP and functioning scales, and lower scores on symptom scales indicate better outcomes.
<sup>3</sup> A HADS total score > 7 indicates a possible anxiety disorder or depression and a score > 11 indicates a probable depression or anxiety disorder.
<sup>4</sup> Higher scores indicate more severe feelings of loneliness.
<sup>+</sup> MD differs from difference mean scores due to rounding.

### Comparison of PROs before and during COVID-19

For 1022 responders (97%) pre and post COVID-19 EORTC scores, and for 942 (90%) pre and post COVID-19 HADS scores could be compared. Overall, mean scores for the EORTC subdomains QoL, physical functioning, role functioning, significantly improved during COVID-19. Mean scores for the EORTC subdomain emotional functioning worsened significantly. Also, median HADS total score and depression score deteriorated significantly during COVID-19 (Table 3).

In the subgroup of actively treated patients, there was a strong significant drop in social functioning of 15·9 points during COVID-19 after adjustment for age, chemotherapy, type of radiotherapy and type of surgery in mixed model analysis (Table 4).

**Table 4.**
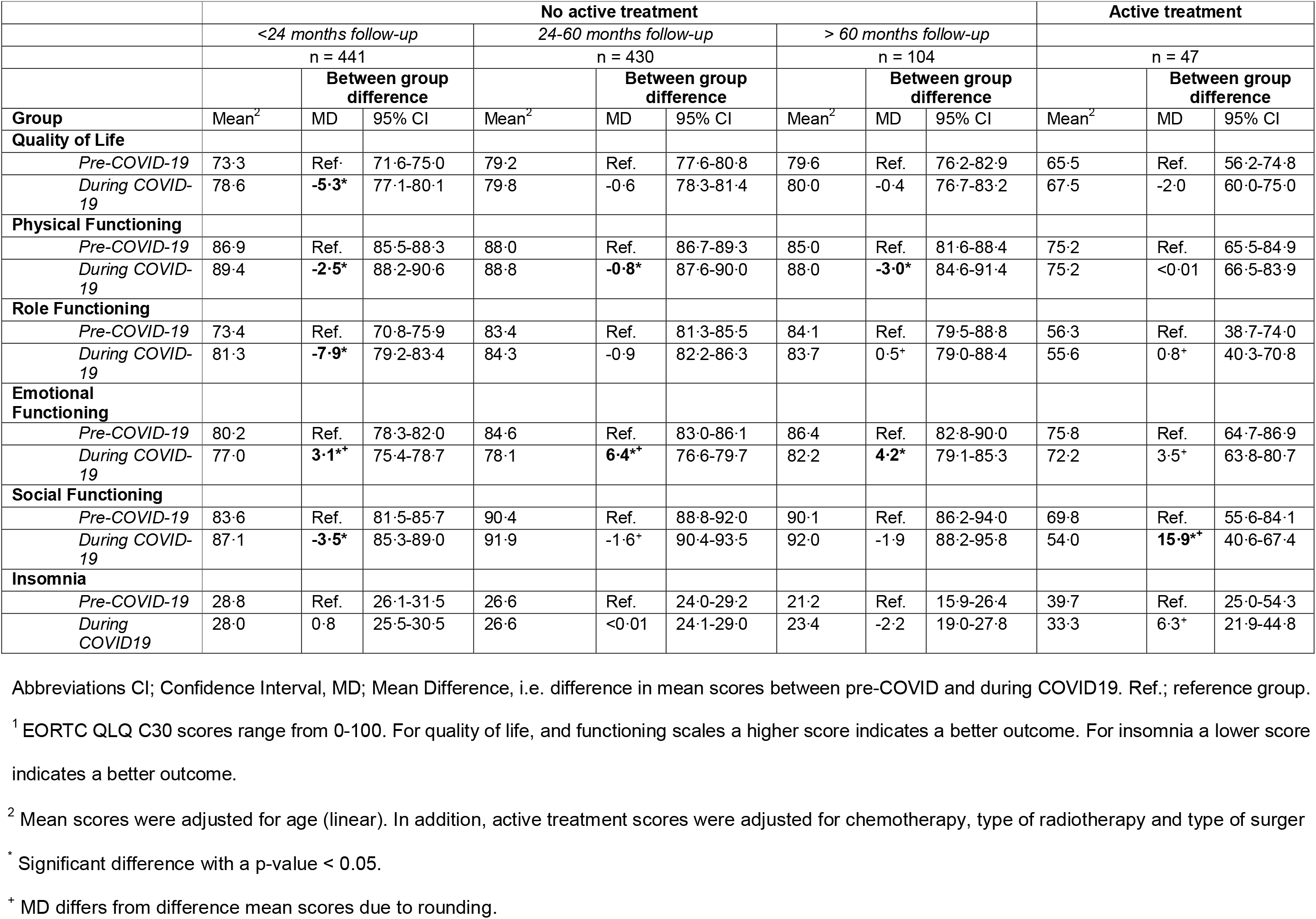
Mixed model analyses in complete cases. The effect of COVID-19 on different EORTC^1^ subdomains of quality of life per follow-up period.

Among the non-actively treated patients, age adjusted analyses showed that emotional functioning worsened significantly in all groups, whereas physical functioning improved significantly in all groups (Table 4). QoL, role functioning, and social functioning improved significantly in non-actively treated patients who were enrolled in UMBRELLA < 24 months (Table 4).

## DISCUSSION

The COVID-19 pandemic has a substantial impact on individuals (being) treated for breast cancer. One in three patients reported to be less likely to contact their general practitioner, and 15% indicated to be less likely to contact their breast cancer physician due to barriers induced by COVID-19 restrictions. In patients actively receiving treatment, social functioning decreased dramatically, and in patients who were no longer receiving active treatment, deterioration of emotional functioning was observed. At the same time, COVID-19 seemed to have a positive effect on QoL, physical functioning, role functioning and social functioning in non-actively treated patients. Loneliness was reported by almost 50% of all participants.

The high proportion of participants indicating to experience a higher barrier to contact their health care providers is in line with the upsetting findings of the Dutch nationwide cancer registry (NKR), who reported a nationwide decrease up to 40% in cancer diagnoses during COVID-19.^26^ Jones and colleagues^27^ from the United Kingdom also expressed their concerns about patients potentially feeling a higher barrier to consult a general practitioner for non-specific symptoms and for moral arguments.^12^ Moreover, an average drop of 37% of referrals by general practitioners to all medical specialties was observed in the Netherlands during the outbreak.^28^ This highlights the importance of creating public awareness about the risk a potential delay in seeking medical help could cause, aiming to lower the barriers for patients to contact a physician when they experience symptoms.^26^

Among patients who were receiving active breast cancer treatment during the COVID-19 pandemic, a major decrease in social functioning was observed (Table 4). One explanation for this decrease could be that these patients were more careful regarding social interaction. Recent publications underlining the risk of COVID-19 related adverse events in cancer patients might have amplified their concerns about contracting the virus.^8,12,29-31^

All non-actively treated patients (i.e., including all lengths of follow-up since diagnosis) showed a significant reduction in the emotional functioning domain. The 4-item emotional functioning domain assesses anxiety, depression and general distress through questions about feeling tense, worrying, depressive, and irritable feelings.^32^ The reduction in this domain is very likely attributable to COVID-19, since we know from pre-COVID-19 work in UMBRELLA that emotional functioning of people treated for breast cancer continues to increase over time (as shown in their supplementary data).^33^ Also, the median score for depression worsened significantly during COVID-19. Concerns about the new viral threat might have enhanced overall uncertainty in individuals. Different types of coping mechanisms could play a role here; lower tolerance of uncertainty is related to higher appraisal of a health threat and higher levels emotion-focused coping strategies.^13^ A previous study showed that, during the 2009 H1N1 viral outbreak, emotion-focused coping was related with increased levels of depression.^13^

Interestingly, despite the deterioration in emotional functioning in all non-actively treated patients, there was a significant increase in global QoL, role functioning, social functioning, and physical functioning. This may partly be explained by the fact that these scores tend to increase over time since diagnosis and, also in the absence of COVID-19, we would expect an increase in these domains.^33^ Another explanation could be found in an effect of a Dutch media campaign that encourages (non-risk) exercise to enhance both public physical and mental wellbeing in these exceptional times.^34^ The remarkable, perhaps counter-intuitive, increase in QoL could be explained by the fact that a shared crisis may put patients’ perceived QoL in relation to their disease in a different perspective, and may even accelerate reconceptualization of their QoL.^35^ The significant increase in social and role functioning suggests that patients reported an increased ability to fulfil responsibilities associated with occupational, and/ or family roles. Governmental measures encouraging work from home and prohibiting social events (i.e., less social obligations) in times of social distancing or lockdown may also play a role.

The current study showed that 48% of all responders felt lonely, the majority reporting to feel emotionally lonely. Experiencing health problems can induce loneliness and vice versa.^36^ Therefore, (breast) cancer patients might be particularly vulnerable for feelings of loneliness. Social isolation measures to fight the COVID-19 pandemic might enhance these feelings. Unfortunately, loneliness is not a parameter that was captured routinely in the UMBRELLA cohort, so it was impossible to measure the impact of COVID-19 on loneliness. However, the reported proportion of 48% loneliness is substantially higher than the reported 34% in the general Dutch population in 2019 (pre COVID-19), as measured by the same Loneliness scale in a survey conducted by Statistics Netherlands (CBS, n = 7.398).^37^ Also, our proportion of patients feeling lonely was substantially higher than the 30-35% cancer patients reporting to feel lonely in a study that was conducted before COVID-19.^38^ Moreover, the percentage of patients feeling severely lonely was strikingly high in this study when compared to other studies; almost 10% compared to 0-2% in other studies performed among cancer patients.^36^ Thus, even though we cannot rule out other contributing factors, the higher proportion of patients feeling lonely in this study is likely due to the COVID-19 measures.

With the high survival rates of breast cancer patients, mental health has become an integral focus of supportive treatment. However, a barrier to e-mental health still exists.^10,39^ Especially in times when face-to-face contact is not an option, and when the global need for psychological and/ or peer support is rising due to the viral threat, efforts are needed to rapidly implement e-mental health programs and digital psychological interventions. Only by adapting to the new circumstances will we be able to treat both ongoing and emerging mental health care conditions due to COVID-19, and prevent long-term problems.^10,40^ The results of this study underline the magnitude of the impact of a major health crisis on the psychosocial wellbeing of breast cancer patients. Considering that a second wave of COVID-19 or another future outbreak with similar impact is probable,^12,41^ one would hope that the current COVID-19 pandemic may serve as a turning point in the adoption and acceptance of successful e-mental health applications.^39^

A limitation of this study is the fact that only 51 patients (4·9%) in our cohort received active treatment during COVID-19, causing relatively wide 95% confidence intervals in this group. As a consequence, the results of this study are predominantly influenced by the large group of breast cancer survivors, while the impact of COVID-19 might be more severe for newly diagnosed patients who, for example, experienced adjusted treatment protocols such as deferred surgery. Second, even though baseline characteristics of responders and non-responders were comparable, an under- or overestimation of the results due to selective (non-)response could not be ruled out as the reasons for 34% non-response were unclear. Third, this study measured the impact of COVID-19 approximately six weeks after the start of COVID-19. Therefore, it is unclear whether the results of this study represent a short-term, or a longer-lasting effect. However, previous literature on the 2009 H1N1 viral threat showed that the psychological effects can persist up to 30 months after the outbreak.^13^ Last, in this study, we could not compare the impact of COVID-19 on patients (being) treated for breast cancer to the impact on a healthy reference population. An important strength of this study is that the UMBRELLA cohort provided a unique opportunity to longitudinally compare validated PRO scores during COVID-19 with the scores before COVID-19 in an identical population, in a representative population of patients with breast cancer.^20^

## CONCLUSION

COVID-19 is having a substantial impact on individuals (being) treated for breast cancer. Emotional functioning deteriorated in patients treated for breast cancer following the COVID-19 pandemic, one in two responders reported (severe) loneliness, and one in three reported to be less likely to contact their health care providers. In actively treated patients, social functioning decreased dramatically. COVID-19 seemed to have a small but significant positive effect on QoL, physical functioning, social functioning, and role functioning in previously treated breast cancer patients.

### Compliance with ethical standards

#### Ethical approval

This study was in accordance with the ethical standards of the institutional and/ or national research committee and with the 1964 Helsinki declaration and its later amendments or comparable ethical standards.

#### Informed consent

Informed consent from all individual participants was obtained within the UMBRELLA cohort.

## Author contributions

The corresponding author (HM Verkooijen) confirms that she had full access to all the data in the study and had final responsibility for the decision to submit for publication.

Each author has contributed significantly to, and is willing to take public responsibility for, the following aspects of the study:

- *Design:* C.A. Bargon, M.C.T. Batenburg, L.E. van Stam, D.R. Mink van der Molen, W. Maarse, N. Vermulst, I.E. van Dam, F. van der Leij, E.J.P. Schoenmaeckers, M.F. Ernst, I.O. Baas, T. van Dalen, R. Bijlsma, D.A. Young-Afat, A. Doeksen, H.M. Verkooijen.
- *Data Acquisition*: C.A. Bargon, M.C.T. Batenburg, L.E. van Stam, D.R. Mink van der Molen, A. Doeksen, H.M. Verkooijen.
- *Analyses*: C.A. Bargon, M.C.T. Batenburg, L.E. van Stam, D.R. Mink van der Molen, D.A. Young-Afat, H.M. Verkooijen.
- *Interpretation* C.A. Bargon, M.C.T. Batenburg, D.R. Mink van der Molen, D.A. Young-Afat, H.M. Verkooijen.
- *Drafting:* C.A. Bargon, L.E. van Stam,
- *Critical Revision*: C.A. Bargon, M.C.T. Batenburg, L.E. van Stam, D.R. Mink van der Molen, W. Maarse, N. Vermulst, I.E. van Dam, F. van der Leij, E.J.P. Schoenmaeckers, M.F. Ernst, I.O. Baas, T. van Dalen, R. Bijlsma, D.A. Young-Afat, A. Doeksen, H.M. Verkooijen.

### Role of medical writer or editor

The authors did not receive any writing assistance.

## Data Availability

The data that support the findings of this study are available from the corresponding author, upon reasonable request.

## Acknowledgements

We would like to thank Janet van Dasselaar for her support with the clinical data management, and Rosalie van den Boogaard for support with IRB approval. We are greatly indebted to the Medical Ethics Committee of the UMC Utrecht for expedited ethical review of the protocol.

## ABBREVIATIONS

BMI: Body Mass Index
CF: Cognitive functioning (EORTC-QLQ30 subdomain)
CI: 95% Confidence Interval
COVID-19: COronaVIrus Disease 2019
DCIS: Ductal carcinoma in situ
EF: Emotional functioning (EORTC-QLQ30 subdomain)
EORTC: European Organization for Research and Treatment of Cancer
HADS: Hospital Anxiety and Depression Scale
IQR: Interquartile range
MD: Mean Difference
METC: Medical Ethics Research Committee (Dutch: Medisch Ethische Toetsingscommissie)
n: number
NKR: Netherlands Cancer Registry
PF: Physical functioning (EORTC-QLQ30 subdomain)
PRO(s): Patient reported outcome(s)
QoL: Quality of Life
RF: Role functioning (EORTC-QLQ30 subdomain)
SD: Standard deviation
SF: Social functioning (EORTC-QLQ30 subdomain)
SPSS: Statistical Package for Social Sciences
STROBE: STrengthening the Reporting of OBservational studies in Epidemiology
UMBRELLA: Utrecht cohort for Multiple BREast cancer intervention studies and Long-term evaluation
UMCU: University Medical Center Utrecht
WHO: World Health Organization

## APPENDIX 1

The following (summarized) advice regarding surgical breast cancer treatment was published by the Dutch Society for Surgical Oncology (NVCO) and the Dutch Society for Gastrointestinal Surgery (NVGIC) on March 27, 2020.^42^

### General advice

1. Based on foreign experiences with COVID-19 patients, sometimes asymptomatic patients appeared to suffer from severe pneumonia. Therefore, one could consider to perform a CT-scan of the thorax shortly before the scheduled surgery in all patients who need intraoperative intubation.

### Breast cancer specific advice

1. Triple negative patients with locally advanced breast cancer who show little response to neo-adjuvant chemotherapy (NAC) should be operated with priority.
2. Surgery should be postponed on the short-term for patients with Her2Neu positive patients with a good response to NAC and who still receive monotherapy with trastuzumab, as well as surgery for hormone receptor positive (HR+) patients without progression under NAC who can already start with hormone therapy (HT).
3. Surgery for patients with only ductal carcinoma in situ (DCIS) and for HR+ patients who can start with HT in the waiting time to their surgery should be postponed for the long-term.
4. It should be considered to perform surgery on patients with positive lymph node(s) (N+) and/ or smaller tumors in order to enable postponement of chemotherapy.

